# Modeling the COVID-19 outbreaks and the effectiveness of the containment measures adopted across countries

**DOI:** 10.1101/2020.04.02.20046375

**Authors:** Edward De Brouwer, Daniele Raimondi, Yves Moreau

**Author notes:** **Disclaimer** The presented results are preliminary and have not been peer-reviewed.

## Abstract

On March 11, 2020, the World Health Organization declared the COVID-19 outbreak, originally started in China, a global pandemic. Since then, the outbreak has indeed spread across all continents, threatening the public health of numerous countries. Although the Case Fatality Rate (CFR) of COVID-19 is relatively low when optimal level of healthcare is granted to the patients, the high percentage of severe cases developing severe pneumonia and thus requiring respiratory support is worryingly high, and could lead to a rapid saturation of Intensive Care Units (ICUs). To overcome this risk, most countries enacted COVID-19 containment measures. In this study, we use a Bayesian SEIR epidemiological model to perform a parametric regression over the COVID-19 outbreaks data in China, Italy, Belgium, and Spain, and estimate the effect of the containment measures on the basic reproduction ratio *R*_0_.

We find that the effect of these measures is detectable, but tends to be gradual, and that a progressive strengthening of these measures usually reduces the *R*_0_ below 1, granting a decay of the outbreak. We also discuss the biases and inconsistencies present in the publicly available data on COVID-19 cases, providing an estimate for the actual number of cases in Italy on March 12, 2020. Lastly, despite the data and model’s limitations, we argue that the idea of “flattening the curve” to reach herd immunity is likely to be unfeasible.

## Introduction

More than 100 countries (1) in the world are currently affected by the coronavirus disease (COVID-19) pandemic (2). COVID-19 is a respiratory infectious disease caused by the SARS-CoV-2 virus (previously known as 2019-nCOV), and it originated in December 2019 in Wuhan (China), most probably following a zoonotic event (3, 4). COVID-19 epidemics are now affecting many European countries, which are at different stages of contagion and containment measures (4). The virus can be found in the respiratory tract of patients 1-2 days before the onset of symptoms, where it shows active replication (5), persisting 7-15 days (4).

Italy was the first to be seriously affected (6), with Spain, France, Belgium, and other countries being 7-14 days behind. Although definitive data on the COVID-19 Case Fatality Rate (CFR) are still missing and the current ones are biased by the testing policies and the demographic structure of the population, the *observed* CFR may be as high as of 10.0% in Italy, 4.0% in China, 6.0% in Spain, and 4.3% worldwide. In Italy, it has been observed that 7-11% of the cases present Acute Respiratory Distress syndrome (ARDS) caused by SARS-CoV-2 pneumonia, and thus require respiratory support in Intensive Care Units (ICUs) (6, 7). European Countries tend to have between 4.2 (Portugal) and 29.2 (Germany) ICU beds per 100,000 inhabitants (8) (EU average is 11.5). This indicates that an exponential-like growth of the COVID-19 cases can rapidly reach oversaturation of the available ICU beds, thereby decreasing the quality of the medical treatments provided to patients and worsening the case fatality rate (6, 9). To avoid this scenario, almost every country affected by the COVID-19 pandemic has put in place measures to contain the epidemic, limiting travels and minimizing physical social interactions, in an attempt to relieve the strain on the healthcare system, in particular ICU units. When it comes to epidemic modeling, these measures affect the *basic reproduction ratio R*_0_, which is typically interpreted as the expected number of cases directly generated by an infected individual in a population susceptible to infection (10). Current estimates of this value range from 2 and 6.5 in China (11–14) and 3.1 in the first phase of the outbreak in Italy (15).

In SEIR (Susceptible, Exposed, Infected, Removed) (16) modeling of epidemics, *R*_0_ = *β/γ*, with *β* representing the number of contacts from an infected individual per unit of time and *γ*^−1^ the period in which a patient is infectious. When *R*_0_ > 1, the number of cases is growing, else, the epidemic is receding. Countries affected by COVID-19 epidemics deployed containment measures that acted on these two parameters. China, for example acted on *R*_0_ by quarantining or hospitalizing cases as soon as they were becoming symptomatic, with an average time elapsed between symptoms and hospitalization of 2.3 (12, 13) or 2.9 days (14). At the same time, China instituted incremental forms of quarantine in Wuhan and the Hubei province, reducing the average number of physical social contacts between residents of the most affected zones, and introduced the use of self-prophylaxis praxis to further reduce the risk of infection, thus effectively reducing *β*.

Similar measures have been adopted by the European countries in which COVID-19 started spreading (4). Italy was the first country affected in Europe, and the first COVID-19 cluster prompted the lockdown of the town of Codogno and later of the Lodi province. The further spreading of the cases led to the lockdown of the most affected regions in northern Italy (Lombardia, Veneto, and Emilia-Romagna) and eventually of the entire country on March 10, 2020. The growth of cases in Spain started with a delay with respect to Italy, but the worrying trend prompted the government to lock down first Madrid and then the entire country. Similar incremental actions have been adopted in Belgium, on the March 13, 2020 and on March 18, 2020.

In this study, we collected the publicly available data regarding cases, recovered and deaths related to the COVID-19 epidemics in China, Italy, Belgium and Spain and we trained a Bayesian SEIR model to perform a parametric regression on these time series. This approach allowed us to model the outbreak progression in those countries inferring the change of basic reproduction ratio *R*_0_ due to the introduction of government-issued containment measures aimed at slowing the outbreak. To do so we used a Markov Chain Monte Carlo (MCMC) approach to fit the SEIR model on the cumulative cases time series by inferring a *β*_*i*_ value corresponding to each containment measure adopted, thus estimating their effectiveness in reducing the transmission or SARS-CoV-2. This approach could help governments *nowcasting* the behavior of the outbreaks and detecting flaws in the containment measures in place and thus act as rapidly as possible, ensuring a proper containment of the disease.

We show that the parameters learned by the SEIR model suggest an *gradual* effectiveness of the containment, with the most drastic effect observed in Spain, with a 71% reduction of *R*_0_ after the measures introduced on March 3, 2020.

We also provide an estimation of the actual number of COVID-19 cases in Italy for March 12, 2020, suggesting that this number have been at that time around 3 times higher than the official cases count.

Finally, despite our model’s limitations, we argue that the idea of “flattening the curve” (i.e., reducing the *R*_0_ of the epidemic to a level that would allow the gradual build up of natural immunity in the population) is likely to be unfeasible. Indeed, reaching herd immunity at a manageable pace is probably not possible in a reasonable time scale.

## Results

### Containment measures in China

We performed a parametric Bayesian regression (see Methods) on the mainland China COVID-19 epidemic data by training a SEIR model on the cumulative cases time series, with the goal of inferring the change in *R*_0_ = *β/γ* produced by the increasingly stringent containment measures introduced by the Chinese government. Such lockdowns and quarantines mainly aim at reducing the frequency of the contacts *β*^−1^ between individuals. We thus used the *β*_*i*_ before and after the introduction of each containment measures and the *γ* as trainable parameters. In this study we used an average incubation time *δ*^−1^ = 5.2 days, as reported in (12).

The implementation of the first containment measure in China happened on February 23, 2020, when all public transportation was suspended in Wuhan, corresponds to a 66% decrease in the inferred *R*_0_, bringing it down from 3.36 (CI 95% [2.88,4.29]) to 1.15 (CI 95% [0.92,1.4]). The introduction of the more stringent measures on February 23, 2020, including closing all non-essential companies and manufacturing plants in Hubei province corresponded to a further reduction to the *R*_0_ identified by our SEIR model, down to 0.19.

### Containment measures in Italy

Italy is the first European country that has been severely hit by the COVID-19 pandemic, and, at the time of writing, it is the second nation in the world in terms of cases, with 92,472. The Italian government reacted to the epidemic by closing all schools and universities on March 4, 2020, putting the north of Italy under lockdown on March 8, 2020, and extending this lockdown to the entire country few days later, on March 10, 2020. On March 20, 2020, the government introduced even stricter measures, banning open-air sports and closing parks and public green.

We used the data of the Italian COVID-19 outbreak provided by the *Protezione Civile* to infer the *R*_0_ before and after the containment measures were implemented. The free parameters are the *β*_*i*_ associated to each containment measure introduced and *γ* that was kept constant across the containment measures.

From this analysis, shown in Fig. 2, it appears that the initially inferred *R*_0_ = 2.68 (CI 95% [2.42, 3.08]) is in line with the 3.1 estimate provided in (15). The *R*_0_ = 1.86 (CI 95% [1.36, 2.37]) inferred after the nationwide lockdown in effect from March 10, 2020 suggests that the effectiveness of the containment measures was gradual and did not immediately bring *R*_0_ below 1. Nevertheless, the data clearly departed from a situation without measures put in place (see Suppl. Fig. 8). When we modeled also the measures introduced on March 20, 2020, the inferred *R*_0_ decreased to 0.46 (CI 95% [0.08, 0.85]), initiating the decrease of the new cases.

**Fig. 1.**
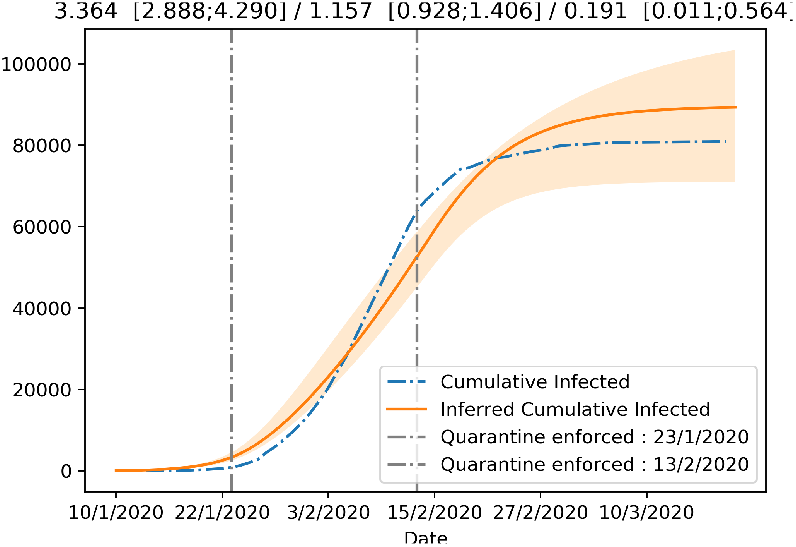
Figure showing the fit of the SEIR model with *β* allowed to change after the introduction of the increasingly strict lockdown measures in China.

**Fig. 2.**
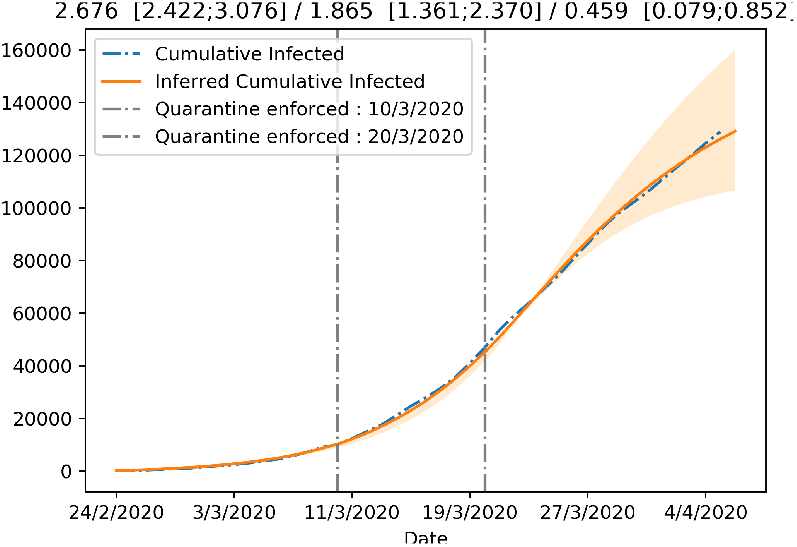
Figure showing the fit of the SEIR model with *β* allowed to change after the introduction of the increasingly strict containment measures in Italy.

### Containment measures in Belgium

Belgium is one of the European countries in which the COVID-19 pandemic apparently arrived later, with the first confirmed case reported on February 4, 2020. The growth of the number of cases in Belgium started from March 1, 2020, and on March 12, 2020, the Belgian government issued containment measures involving the closure of schools, cafes and restaurants starting from March 14, 2020. The government then extended these measures, enforcing stricter “physical distancing”, starting from March 18, 2020, at noon.

In Fig. 4, we can see that the SEIR model infers a change in *R*_0_ that increases it from the original 2.1 (CI 95% [1.94,2.12]) to 3.19 (CI 95% [2.43,4.14]). The fact that the lockdown leads to an increase of *R*_0_ is very surprising and could be explained by the number of tests carried out during that period (see Discussion).

**Fig. 3.**
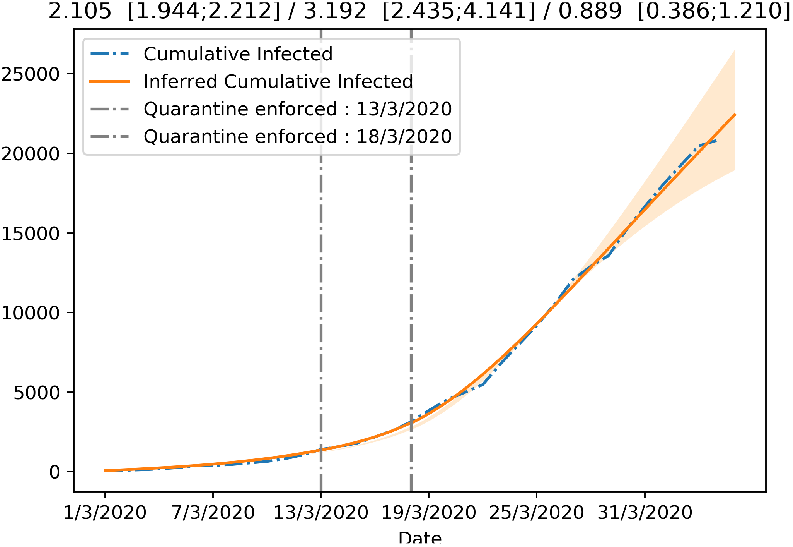
Figure showing the fit of the SEIR model with *β* allowed to change after the introduction of the increasingly strict lockdown measures in Belgium.

**Fig. 4.**
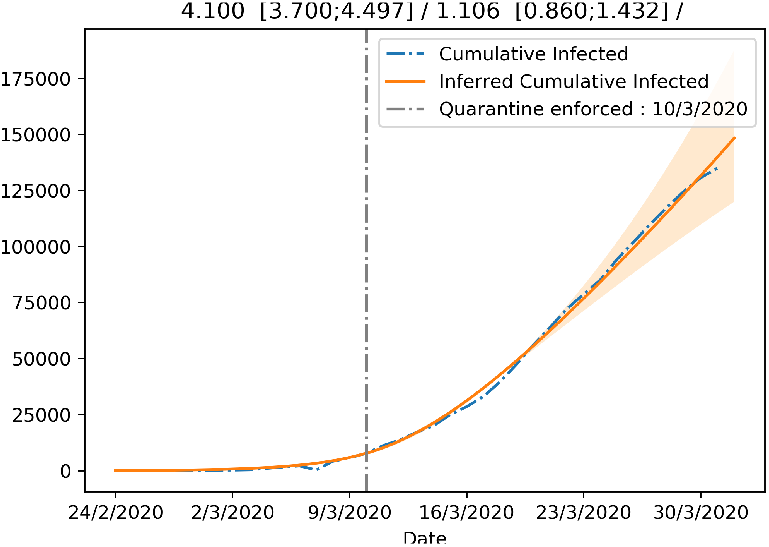
Figure showing the fit of the SEIR model with *β* allowed to change after the introduction of the increasingly strict lockdown measures in Spain.

The second set of measures appears to be more effective with a reduction of *R*_0_ to 0.88 (CI 95% [0.39,1.21]), initiating the decrease of the new cases.

### Containment measures in Spain

Although the first COVID-19 case in Spain dates to March 1, 2020, the epidemic there did not show worrying numbers until the end of February, with a rapid growth starting from the beginning of March. This crisis was answered first with containment measures in the Community of Madrid, enforced from March 11, 2020, followed by nationwide measures enforced from March 15, 2020.

We fitted the SEIR model to infer the change in *β* resulting from the implementation of the containment measures. Similarly to the Italian and Belgian case, we observe a clear decrease in *R*_0_, but the value inferred by the model is still slightly above 1, indicating a significantly reduction in the acceleration of the epidemic, but still insufficient to revert the increase of the new cases. This might be due to the effect of under-reporting of the actual number of COVID-19 cases on our SEIR model.

### Analysis of reporting in official counts and biases in testing

In an attempt to address the under reporting of cases, we computed a reasonable estimate of the actual number of COVID-19 cases in Italy on March 12, 2020. To do so we relied on the fatality rate (CFR) of the disease and the age distribution of the cases in South Korea, which adopted an extensive testing strategy to face the COVID-19 crisis, administering one test every 142 citizens. Since South Korea has tested a very large part of its population with no evident biases, we considered this to be the most reliable data when it comes to reporting the actual numbers and age group of infected individuals. South Korea shows indeed a Pearson correlation coefficient between the number of cases detected among 10-yrs age bins and its demographic structure of *r* = 0.69 (p-value= 0.039), while Italy has an *r* = 0.21 (p-value= 0.591), suggesting a much more skewed testing. Our estimation is based on three other assumptions. First, we assume that the disease propagated similarly in South Korea and Italy over the different age bins. Second, we assume that South Korean and Italian healthcare have similar standards, thus suggesting a comparable fatality rate once the testing bias is addressed. Third, we assume that the healthcare system in Italy (e.g., the availability of ICU beds) has not reached saturation, and to satisfy this condition we indeed chose to perform this estimation for March 12, 2020, as lockdown measures in Italy appear to be the result of the healthcare system rapidly approaching saturation.

We first adjusted the South Korean number of cases by age group with respect to the demographic structure of the Italian population. As reported on Figure 5 (green bars), the age of confirmed patients is heavily skewed towards older individuals in Italy, while it is more consistent with the demographic structure in the South Korean data. We argue that the skewness of the Italian cases towards older age groups results from the fact that on the February 26, 2020 on the Italian testing strategy changed from *blanket testing* to focusing on symptomatic and high-risk individuals (15), thus introducing a clear sampling bias. We then adjust the proportion of cases per age in Italy by computing

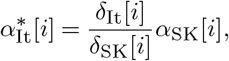

Where 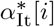 is the non-normalized proportion of Italian cases in age bin *i, α*_SK_[*i*] the proportion of South Korean cases in age bin *i* and *δ*[*i*] are the proportion of the corresponding age bin in the total demographics of the respective country. The normalized proportion of Italian cases *α*[*i*] is obtained by dividing by the sum of *α*^∗^[*i*] over all age bins. They are reported in orange on Figure 5.

**Fig. 5.**
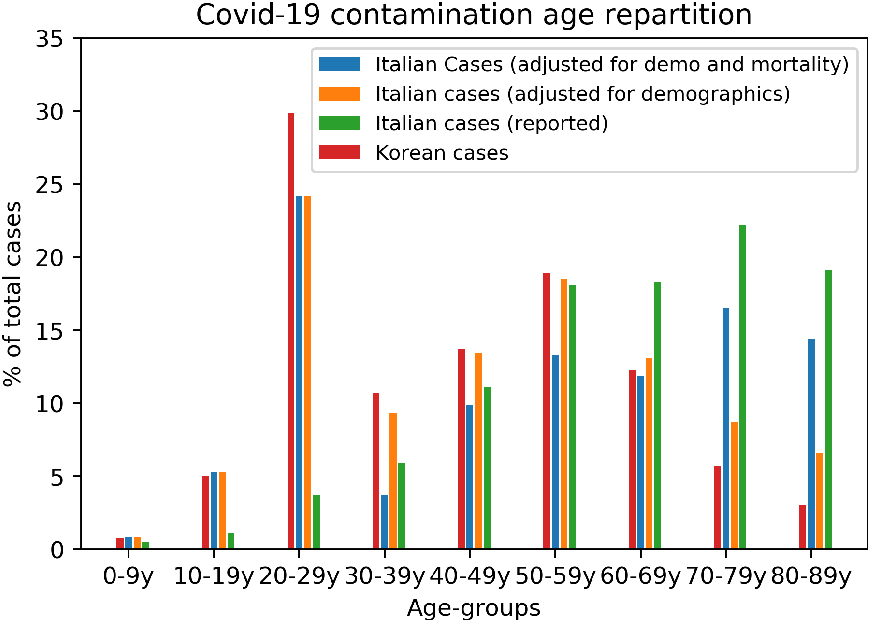
Proportions of COVID-19 cases per age bins in South Korea (Blue) and Italy (Green). The orange bars give the estimated Italian distribution when adjusted for the age sampling bias with demographics only. The red bars additionally accounts for the number of deaths in Italy in each age bin. The data we used was the one available on the March 12, 2020 for Italy and South Korea.

In a second stage, we compute the expected number of cases in Italy based on the number of deaths and use it to further adjust the age distribution. We use South Korea death rates per age bins to infer the number of closed cases. We compute the expected number of cases per age bin by dividing the number of deaths by the fatality rate measured in South Korea:

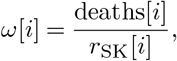

where *ω*[*i*] is the number of Italian closed cases in each age bin *i*, deaths[*i*] is the number of Italian deaths in that bin and *r*_SK_ is the corresponding fatality rate. Applying this formula for *ω* will result in undefined numbers for the youngest age bins. This effect is caused by the very low mortality at those ages (the denominator or the numerator is 0). In our case, bins from 0 to 30 years old were undefined. We address this issue by using the corrected age distribution of Italian cases *α*_It_. We compute the proportions of cases over 30 years old as

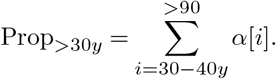

We then compute the estimated total number of closed cases in Italy as

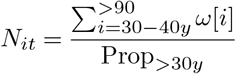

Using the data available on March 12, 2020 for Italy and South Korea, we find an estimated number of *real* cases *N*_*it*_ = 45, 052. This is to be compared with the total number of reported cases in Italy at that date which is 15, 113. The inferred number of cases is thus 3 times higher than the reported figure.

The undefined *ω* bins can then be inferred by multiplying their *α* with the total number of cases *N*_*it*_. The resulting distribution of cases per age is presented on Figure 5 in red.

### Analysis of the healthcare system strain-level during the epidemic progression

The COVID-19 pandemic has been putting immense pressure on the healthcare systems of many countries because it spreads widely in the population in an asymptomatic or mild form (4), but a significant percentage of the symptomatic cases (6-11% in Italy (6, 7)) requires ICU treatment, which is a limited resource in any country, including European countries (8). The availability of ICU beds is crucial (9) because so far there is no established curative treatment for COVID-19 and the clinical best practice is to put patients suffering from Acute Respiratory Distress Syndrome under respiratory support, for a period that may last up to two weeks (6). Saturation of ICU capacity causes a dramatic decrease of the quality of the medical treatments provided to patients, thereby worsening the observed Case Fatality Rate (CFR) (6, 9).

To analyze the burden that the COVID-19 epidemic brings to the national healthcare systems of the affected countries, we plot the evolution over time of the log-CFR (the log-ratio of deaths per confirmed COVID-19 case). More specifically, it is the logarithm in base 10 of the ratio of the cumulative number of deaths over the cumulative number of cases. We used the cumulative numbers as they are more stable that the instantaneous ones.

During the first days of the epidemics, we expect this rate to be noisy during the first phase of the epidemics. Due to the delay between infections and deaths, the number of deaths remains very low during the first days while the number of cases increases exponentially. After this transition period, first deaths occur and if ICU units do not reach saturation, we expect this rate to be stay constant. However, if this rate increases, this suggests that the healthcare system is under strain as it tries to cope with the growing number of patients requiring ARDS treatment. Towards the end of the epidemics, when the cumulative number of cases flattens out, the rate is expected to increase, again due to delay between the reporting of cases and the occurrence of deaths.

Fig. 6 shows this log-ratio over time in China. After some expected initial oscillations (also possibly due to the change in the policies regulating the detection of the cases), China showed a steep increase incidence of deaths with respect to the number of cases, indicating a significant strain over its healthcare system, and possibly the degradation of the quality of the care provided. Towards, the end of the epidemics, when the number of cases flattens, we observe a slower steady increase.

**Fig. 6.**
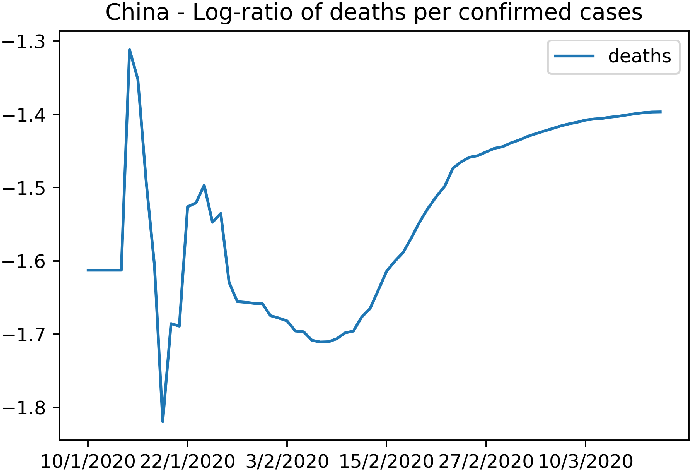
Figure showing the strain that the COVID-19 pandemic put on the NHS of China.

The same plot is shown for Italy in Figure 7. Shortly after the first days of the epidemics, the mortality rate started growing quickly, suggesting an increasing strain on ICUs and the Italian healthcare system, as reported also in (9).

**Fig. 7.**
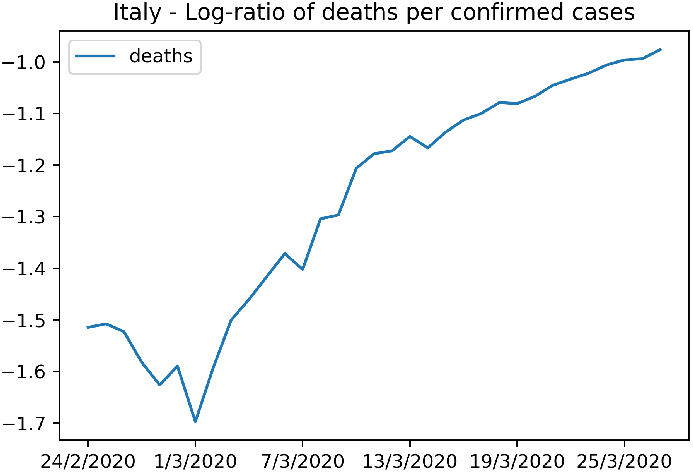
Figure showing the strain on the Italian national health services from the COVID-19 epidemic.

Again, it is important to notice that both number of deaths and cases are likely to be significantly under-reported, although possibly in different proportions and thus impact our conclusions.

## Discussion

### Effectiveness of containment measures

Containment measures in China significantly reduced COVID-19 spreading. The first lockdown resulted in a 65% decrease of the reproduction factor *R*_0_ and the second, stricter wave of measures eventually managed to bring it to close to 0. We do however observe that while the model was able to fit the Italian, Belgian, and Spanish data relatively well, its fit of the data from China was rather mediocre. We are unsure about what could have caused this discrepancy.

Containment measures in Italy appear to have had a more gradual effect. The reason for this is not entirely clear, but data from the Lombardy region, based on anonymous cell phone tracking (not showed here), suggests that almost 40% of the population of Lombardy were still commuting and moving around notwithstanding quarantine measures, although the trend from February 26, 2020 and March 16, 2020 clearly indicates a progressive reduction of displacements. This progressive trend is also consistent with the Community Mobile Reports provided by Google.

Such large percentage of the population moving across Lombardy might in part be explained by the fact that factory closures were only partial until March 23, 2020, when the Italian government issued a decree mandating the immediate halt of all non-essential production, industries, and businesses across the country. Data from the Italian Ministry of Interior (17) also indicates that during the lockdown 1.7 million police controls were carried out with infractions to the containment registered in 4% of controls.

The analysis of the data from Belgium suggests at first that the enforcement of the first lockdown measures were counter-productive. This is very unlikely. Instead, a plausible explanation may lie in the variation of number of tests during the the transition period between the two belgian lockdowns. As show on Figure 10 in the Supplementary, the number of tests significantly dropped during this period, leading to under-reporting of new infections. When, 5 days later, the number of tests increases again, so does the number of newly infected patients. Under the assumptions of our model, this rapid but rather artificial sudden increase of infected patients is supposed to have been contaminated by patients infectious during the between lockdowns period. As this number was underestimated, the *R*_0_ automatically increases to account for the too low number of infectious patients.

More generally, the compliance of the population to the measures was progressive as suggested by seismic data (18) and community mobility reports of Google.

### Data biases and heterogeneity in the testing strategies limit the ability to draw clear conclusions

The models fits presented in the Results section are based on the officially available COVID-19 cases counts from China, Italy, Belgium, and Spain. Even from a superficial analysis of this data, several biases that hinder the modeling of these outbreaks become clear.

First, the number of tests that can be run each day is finite, because of the limited availability of supplies and personnel, making *blanket testing* currently impossible to perform in many countries. This results in a large number of unreported cases with respect to the available data.

Second, if tests are performed mainly on symptomatic patients for diagnostic purposes, because of the generally higher age of the hospitalized cases, the resulting official COVID-19 cases data will show a striking proportion of patients over 60 years old, regardless of the actual demographic structure of the population (see Suppl. Fig. 9).

Another reason why the sheer number of tests performed is not a clear indication of the level of bias present in the data is that the number of test performed is just an upper bound for the actual number of individuals screened, because for example medical personnel with high risk of exposure may undergo periodic tests. Moreover, the directives of the Italian Ministry of Health indicates that a COVID-19 patient must be negative to two consecutive tests performed with a 24h delay (19) to be considered as having recovered from the disease.

The cumulative number of cases we used to fit the SEIR model is therefore most probably both severely under-estimated and skewed towards older age groups in the population. Both in Belgium and in Italy, for instance, patients who are diagnosed as suspect COVID-19 case over the phone by their GP, but who present no immediate risk of complication, are nor tested, nor reported as new cases. As the epidemic progresses and healthcare resources become mobilized, testing capacity increases and we observe a growing number of newly tested individuals.

Yet a key assumption of our model is that new infections are caused by contamination from currently reported infectious individuals, because our modeling is based on the *observed* cases, for which official data exists. However, in practice, many of the newly diagnosed patients have been infected by the majority of unreported infectious people. Our model will thus infer an higher *R*_0_ to compensate for the underestimated pool of infectious patients. This might explain the seemingly high values of *R*_0_ estimates in the inter-lockdowns period in Belgium for instance.

Interestingly, the estimation of the actual (vs. reported) number of cases on March 12, 2020 in Italy suggests that, although heavily under-represented in the official data because of testing bias, the 20-29 age group is the most affected by COVID-19. Given that age group is particularly socially active, one might speculate that infections via this age group may have played a key role in the spread of COVID-19 across Italy, even though these cases ended up almost completely unreported. There are however some limitations to this analysis. While South Korea’s testing strategy has clearly been comprehensive, it is not clear that it has been completely unbiased. In particular, the low number of cases in the 10-19 years bin compared to the 20-29 years bin might be explained by a radical difference in the true proportion of cases between those two age groups, but also by lower testing among younger individuals because they might have been considered at very low risk of complications and/or unlikely to be infectious. The information available does not allow us to discriminate between those explanations. Moreover, the assumption that the case fatality rate in South Korea and (north) Italy is identical is subject to discussion as physicians in South Korea are more experienced in managing patients suffering from Acute Respiratory Distress Syndrome following the 2015 Middle East Respiratory Syndrome (MERS) epidemic in South Korea.

Moreover, every country adopted its own specific strategy for testing and reporting of cases, resulting in heterogeneity of the COVID-19 data coming from different countries. For example, South Korea opted for blanket testing of its population and selective quarantine of the positive cases, while Italy focused on testing high-risk and symptomatic subjects and generalized lockdown of the country to reduce the *R*_0_ by acting on the frequency of social contacts.

Even within the same country, the reporting strategy changed over time in some cases, leaving a trace in the data. For example, the number of daily new cases in China presents an unlikely *spike* of 14,108 new cases in a single day (February 12, 2020) because of a change in the reporting strategy, since also clinically diagnosed COVID-19 cases started to be included in the cases count, alongside laboratory tests. This measure was probably necessary to overcome the saturation of the maximum number of tests that could be performed every day, but caused the sudden inclusion of previous “suspect” cases in the official count. Similarly, Italy opted for testing only high-risk individuals and symptomatic cases from February 26, 2020 on (15).

### Limitations of the model

The SEIR model does make important assumptions and shows significant limitations. The size of the population is considered constant without births and external deaths. Given the time scale for studying the epidemics here, this assumption is likely to have a negligible effect. Next, individuals who have recovered from the disease are considered to be immune. Based on what is known about SARS-CoV-1 and SARS-CoV-2, which are closely phylogenetically related, it is reasonable to assume that individuals who recover from COVID-19 will benefit from immunity at least over the period modeled here. Also, it has been reported that asymptomatic and mildly symptomatic individuals (who will not be recognized as carrying the disease), and presymptomatic individuals (during at least 2-3 days of the incubation period) are likely to be contagious with a degree of infectivity that is not yet well characterized. The SEIR model does not account for these effects and the high value of *β* obtained might in part be caused by the need to account for those missing contagion events. Moreover, the number of symptomatic individuals might also be underestimated because testing is in some cases being focused on most severe cases, which similarly will lead to the inflation of *β* and *R*_0_.

More sophisticated models (with more patient compartments) might better capture the different effects described before, but such models will have significantly more free parameters, which means that those parameters might simply be unidentifiable from the available data or that overfitting is likely.

### About “flattening the curve”

Despite their limitations, our models show that the idea of “flattening the curve” (i.e., reducing the *R*_0_ of the epidemic to a level that would allow the gradual build up of natural immunity in the population) is likely to be unfeasible. Any significant reduction of *R*_0_ that would not bring it extremely close to 1 would overwhelm the healthcare system because the ICU capacity and the height of the epidemic peak in a immunologically naïve population are simply on different scales (in the SIR, the proportion of the population infectious at the epidemic peak is given by 1−1*/R*_0_−ln(*R*_0_)*/R*_0_. For example, 30% of the population is infectious at the epidemic peak for *R*_0_ = 3, while the ICU capacity in for example Belgium is 15.9 beds per 100,000 inhabitants (8)). Even if the epidemic could be controlled at a fixed level corresponding to a heavy but non-overloading load of the ICU capacity, the time needed to build herd immunity would be measured in years. As an example, a back-of-the-envelope calculation for Belgium based on a permanent ICU capacity of 1,000 beds for coronavirus patients (compared to the pre-existing capacity of 1,750 (8) beds, which would mean a major continuing strain on the hospital system and thus the need to maintain supplementary capacity for several years), assuming an average ICU stay of 10 days (6, 20), and assuming that 2% of patients affected in the general population would eventually require ICU care, would mean that 100 patients would be admitted at ICU care per day and that 5,000 individuals in the general population would be infected by the disease each day. Reaching a level where 50% of the population (of about 11 million people) has achieved natural immunity would require 1,100 days or 3 years. Given that the immunity to the disease might be relatively short-lived (around 2 years for SARS (21)), it might simply be next to impossible to achieve herd immunity without overwhelming the healthcare system.

Moreover, such a strategy would require maintaining the number of cases in the population at a tightly controlled level with *R*_0_ being maintained on average at 1. Whenever *R*_0_ would be above 1, the disease would flare up, which would quickly overload a healthcare system maintained at saturation. When *R*_0_ would be below 1, the disease would start vanishing, which would extend the time needed to build herd immunity. Given that it is completely unclear what the precise impact of any containment measure is on *R*_0_, a strategy based on lifting and reimposing measures to switch between *R*_0_ slightly below 1 and *R*_0_ slightly above 1 does not appear realistic.

If a treatment became available that would greatly diminish the risk of complications (for example, by a factor 10), or if it turns out that the proportion of the general population that develops severe complications when infected by SARS-CoV-2 is much lower than 2%, it might be possible to revisit strategies based on “flattening the curve”.

In the absence of such a silver bullet treatment, the only plausible option for the moment seems to be the immediate quashing of the epidemic together with the development of strategies to try to contain the disease at a minimal level driven by imported cases, while waiting for greatly improved treatments or a vaccine. In such strategies, as currently deployed by South Korea, Hong Kong, and Singapore for example, patients only arise from imported cases and small local clusters that are rapidly quashed. It is likely that such strategies will sometimes fail in insufficiently prepared populations leading to the reimposing of heavy quarantine measures during the time needed to quash the new epidemic flare. It seems advisable to reimpose strict quarantine measures as soon as uncontrolled local circulation of the disease is suspected.

## Methods

### Data collection

We collected COVID-19 data from official sources. The Italian Protezione Civile releases COVID-19 data daily on its git repository^1^. Chinese COVID-19 data has been obtained from the official bulletins^2^. Belgian COVID-19 data has been collected from Sciensano, the Belgian center for epidemiology of infectious diseases ^3^. The Spanish COVID-19 data has been obtained from the official bulletins^4^. The South Korean data has been collected from the official KCDC press releases^5^.

### SEIR model

The SEIR (Susceptible, Exposed, Infected, Recovered) is a widely used mathematical model for the description of the behavior of infectious disease outbreaks. It consists in the following system of ordinary differential equations (ODEs):

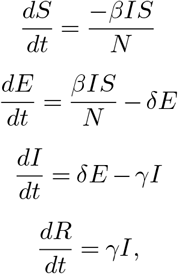

where the variables *S, E, I*, and *R* are respectively the number of (1) susceptible individuals (not immune and never exposed to the virus), (2) exposed individuals (currently incubating the disease), (3) the infectious symptomatic individuals, and (4) recovered (healthy and currently immune) and deceased individuals. The total number of individuals in the population is assumed to remain constant with value *N* = *R*(*t*) + *I*(*t*) + *S*(*t*). We further define *C*(*t*), the cumulative number of cases as *I*(*t*)+ *R*(*t*).

The parameters of this model are *δ, β* and *γ*. Those tune the dynamics of the epidemics. *δ*^−1^ can be interpreted as the average incubation period (i.e., the average time spent in pool *E* before becoming infectious *I*). *β* corresponds to the average number of infections an infectious individual will cause per unit of time and *γ*^−1^ corresponds to the average time necessary to recover from the disease (i.e., going from *I* to *R*). The average number of new infections arising from a single infectious person is then *R*_0_ = *β/γ*. In this study we set *δ*^−1^ = 5.2 days (12) and we used *β* and *γ* as trainable parameters. We inferred a single *γ* for each country, therefore keeping both *γ* and *δ* constant over time, before and after the introduction of containment measures. The effect of these measures is then modelled by a change in *β*. Importantly, in this work, we considered that the measures enforcement resulted in a adaptive change in *β*. This is motivated by the fact that people are only gradually adopting the enforced measures, as suggested by seismic data (18) and mobile data (see Discussion for more details).

We assumed that the beta was then exponentially adapting to a new target. Let *β*_0_ be the *β* right before lockdown enforcement. We posit that the *β* after lockdown is decaying towards its target *β*_∞_ as:

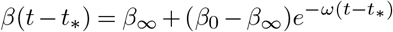

where *t*_∗_ stands for the time when the lockdown was enforced. In this text, we reported the *R*_0_ computed with *β*_∞_ to study the impact of the measures. When 2 or more distinct lockdowns are enforced over time, we take as *β*_0_ the value of *β* at the time of lockdown of interest. We fixed 1*/ω* = 4 days for all countries, meaning that after a week, *β* reached 85% of the gap between its target and its starting value. This value is inline with the sismic and mobile data (18).

We inferred the *β* parameters during each period by fitting the cumulative number of cases *C*(*t*) with MCMC (Metropolis Hastings). We use a Poisson likelihood and uniform priors for *β*, such that *β* ∼ 𝒰 (0, 2), and we use another uniform prior for *γ*, such that *γ* ∼ 𝒰 (0.2, 1). This leads to plausible values of *γ*^−1^ between 1 and 5 days which is inline with the current estimates for the mean time elapsed between onset of symptoms and hospitalization in China (between 2.3 (12, 13) and 2.9 days (14)) and Italy (4days (22)).We inferred the *β* parameters during each period by fitting the cumulative number of cases *C*(*t*) with MCMC (Metropolis Hastings). We use a Poisson likelihood and uniform priors for *β*, such that *β* ∼ 𝒰 (0, 2), and we use another uniform prior for *γ*, such that *γ* ∼ 𝒰 (0.2, 1). This leads to plausible values of *γ*^−1^ between 1 and 5 days which is inline with the current estimates for the mean time elapsed between onset of symptoms and hospitalization in China (between 2.3 (12, 13) and 2.9 days (14)) and Italy (4days (22)).

Additionally, to allow for more flexibility, we set a Gaussian prior on the initial value of the infectious pool *I*(0). The initial values of others pools are taken as the ones reported in the available data. We generated 10,000 samples from the posterior distribution of *β* and discard the first 5,000 as burnin period. For numerical integration, we used Euler with a time delta of 0.05 days.

## Data Availability

Data is publicly available

## ACKNOWLEDGEMENTS

The authors would also like to thank Umberto Rosini and the Protezione Civile Italiana for sharing the COVID-19 outbreak data on github. DR is grateful to Anna Laura Mascagni, Angela Motz, Paolo Rizzi and Marco Simonini for the constructive discussion. EDB is grateful to Giulia Lazzara for the inspirational exchanges. YM is funded by Research Council KU Leuven: C14/18/092 SymBioSys3; CELSA-HIDUCTION CELSA/17/032 Flemish Government:IWT: Exaptation, PhD grants FWO 06260 (Iterative and multi-level methods for Bayesian multirelational factorization with features) This research received funding from the Flemish Government under the “Onderzoeksprogramma Artificiële Intelligentie (AI) Vlaanderen” programme. EU: “MELLODDY” This project has received funding from the Innovative Medicines Initiative 2 Joint Undertaking under grant agreement No 831472. This Joint Undertaking receives support from the European Union’s Horizon 2020 research and innovation programme and EFPIA. DR is funded by a FWO postdoctoral fellowship and EDB is funded by a FWO-SB grant.

## Supplementary Note 1: Supplementary Figures

**Fig. 8.**
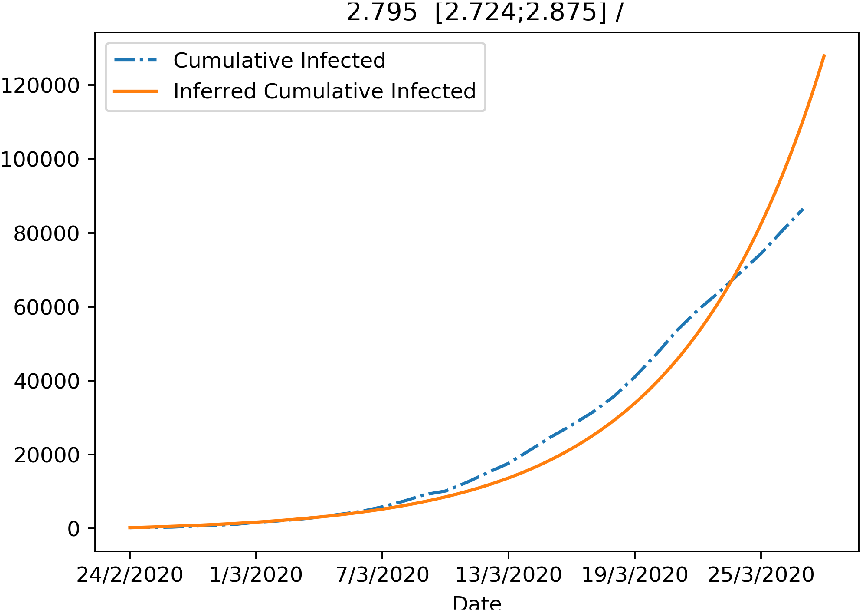
Figure showing the fit of the SEIR model with no Quarantine (no changes in betas allowed).

**Fig. 9.**
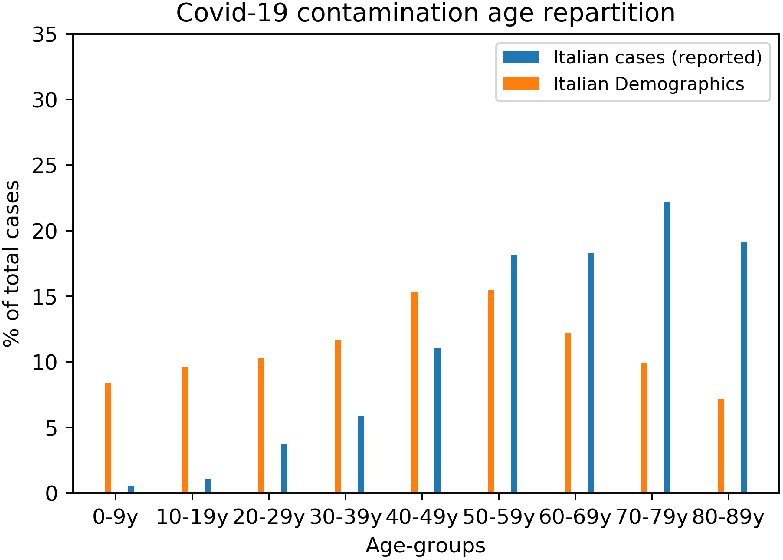
Demographics age repartition of Italy along with the age distribution of reported cases.

**Fig. 10.**
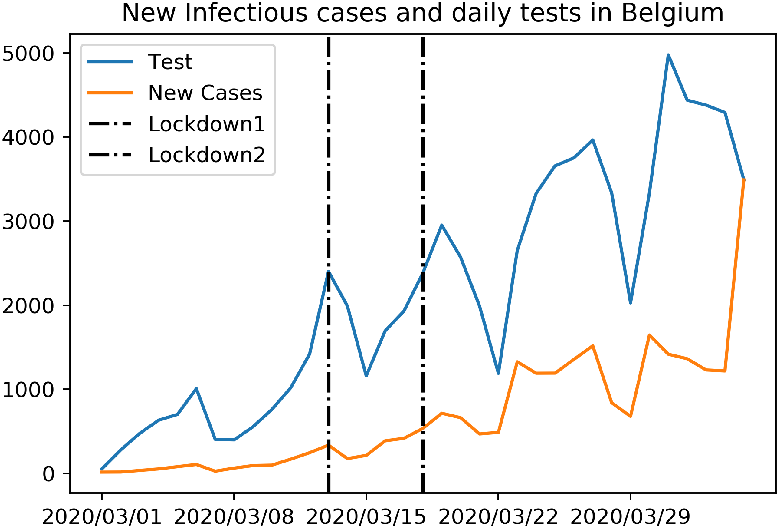
Evolution of tests and new infectious cases in Belgium over time.

## Footnotes

1 https://github.com/pcm-dpc/COVID-19

2 http://www.nhc.gov.cn/xcs/yqtb/list_gzbd.shtml

3 https://epidemio.wiv-isp.be/ID/Pages/2019-nCoV.aspx

4 https://www.mscbs.gob.es/profesionales/saludPublica/ccayes/alertasActual/nCov-China/situacionActual.htm

5 https://www.cdc.go.kr/board/board.es?mid=a30402000000&bid=0030

